# Polygenic risk score-based phenome-wide association study identifies novel associations for Tourette syndrome

**DOI:** 10.1101/2022.09.01.22279340

**Authors:** Pritesh Jain, Tyne Miller-Fleming, Apostolia Topaloudi, Dongmei Yu, Petros Drineas, Marianthi Georgitsi, Zhiyu Yang, Renata Rizzo, Kirsten R. Müller-Vahl, Zeynep Tumer, Nanette Mol Debes, Andreas Hartmann, Christel Depienne, Yulia Worbe, Pablo Mir, Danielle C. Cath, Dorret I. Boomsma, Veit Roessner, Tomasz Wolanczyk, Piotr Janik, Natalia Szejko, Cezary Zekanowski, Csaba Barta, Zsofia Nemoda, Zsanett Tarnok, Joseph D. Buxbaum, Dorothy Grice, Jeffrey Glennon, Hreinn Stefansson, Bastian Hengerer, Noa Benaroya-Milshtein, Francesco Cardona, Tammy Hedderly, Isobel Heyman, Chaim Huyser, Astrid Morer, Norbert Mueller, Alexander Munchau, Kerstin J Plessen, Cesare Porcelli, Susanne Walitza, Anette Schrag, Davide Martino, The EMTICS collaborative group, Andrea Dietrich, The TS-EUROGRAIN Network, Carol A. Mathews, Jeremiah M. Scharf, Pieter J. Hoekstra, Lea K. Davis, Peristera Paschou

**Author notes:** **Corresponding Authors** Dr. Peristera Paschou, Lilly Hall of Life Sciences, Purdue University 915 W. State Street, Room 1-225, West Lafayette, IN 47907., Dr. Lea K Davis, 511-A Light Hall, Vanderbilt University 2215 Garland Ave, Nashville, TN 37232.

## Abstract

Tourette Syndrome (TS) is a complex neurodevelopmental disorder characterized by vocal and motor tics lasting more than a year. It is highly polygenic in nature with both rare and common previously associated variants. Epidemiological studies have shown TS to be correlated with other phenotypes, but large-scale phenome wide analyses in biobank level data have not been performed to date. In this study, we used the summary statistics from the latest meta-analysis of TS to calculate the polygenic risk score (PRS) of individuals in the UK Biobank data and applied a Phenome Wide Association Study (PheWAS) approach to determine the association of disease risk with a wide range of phenotypes. A total of 57 traits were found to be significantly associated with TS polygenic risk, including multiple psychosocial factors and mental health conditions such as anxiety disorder and depression. Additional associations were observed with complex non-psychiatric disorders such as Type 2 diabetes, heart palpitations, and respiratory conditions. Cross-disorder comparisons of phenotypic associations with genetic risk for other childhood-onset disorders (e.g.: attention deficit hyperactivity disorder [ADHD], autism spectrum disorder [ASD], and obsessive-compulsive disorder [OCD]) indicated an overlap in associations between TS and these disorders. ADHD and ASD had a similar direction of effect with TS while OCD had an opposite direction of effect for all traits except mental health factors. Sex-specific PheWAS analysis identified differences in the associations with TS genetic risk between males and females. Type 2 diabetes and heart palpitations were significantly associated with TS risk in males but not in females, whereas diseases of the respiratory system were associated with TS risk in females but not in males. This analysis provides further evidence of shared genetic and phenotypic architecture of different complex disorders.

## Introduction

Tourette Syndrome (TS) is a complex neurodevelopmental disorder characterized by vocal and motor tics lasting more than a year^1^. It is a childhood onset condition with a prevalence of 0.6 – 0.8% in school age children and has a population heritability of 70% and SNP-based heritability estimates ranging from 0.21 to 0.58^2–6^. TS is highly polygenic in nature and multiple genetic variants account for a substantial proportion of phenotypic variance of the condition^6–11^.

Epidemiological studies have shown TS to be associated with various phenotypes and disorders, but large-scale phenome-wide analyses in biobank level data have not been performed to date^12,13^. This is likely due to the scarcity of individuals with TS in biobank-scale available datasets. To address this, polygenic risk scores (PRS), a measure of an individual’s genetic predisposition for a disease, can be used as a proxy for the case-control status of TS^14^. Such scores can be used to identify different general health, mental health, and socio-demographic outcomes associated with the genetic risk of a disorder. They may also help to identify other phenotypes associated with TS genetic risk and explore causal relationships between TS and other phenotypes^15^.

PRS-based phenome-wide association studies (PheWAS) have become an increasingly common method to identify different factors associated with the genetic risk of a complex disease^16–19^. Since the scores are calculated based on genomic information that is fixed at birth, this method has the advantage of being less susceptible to reverse causality^15^. Applying PRS based PheWAS methods across large datasets like the UK Biobank has led to the identification of novel phenotypes associated with genetic risk of psychiatric disorders that have been previously correlated with TS^20,21^, such as autism spectrum disorder (ASD), attention deficit hyperactivity disorder (ADHD), schizophrenia and major depressive disorder^,22^. These associations have also shed light on the shared genotypic architecture of such related disorders.

Here, using the summary statistics from the latest GWAS meta-analysis of TS^11^ and genomic and phenomic data from the UK Biobank (UKB), we conducted a PRS-based PheWAS for TS, interrogating a wide range of phenotypes including physical and mental health, lifestyle, and socio-demographic factors. Our goal was to uncover phenotypes that may be associated with TS genetic risk and compare to phenotypic associations with genetic risk of other disorders along the impulsivity-compulsivity spectrum that have previously been found to be genetically associated with TS^20,21^. This work furthers our knowledge on the TS phenotype and genetic architecture and can serve as a basis for future analysis that will investigate potentially causal relationships between the phenotypes that are found associated with TS genetic risk.

## Methods

### Study Population and Quality control

The UK Biobank dataset which has data from ∼500,000 individuals was used for the PheWAS analysis. First, we removed individuals with non-white British ancestry based on the self-reported ancestry information available in the UK Biobank data. Next, we removed individuals with greater than third degree relatedness based on the kinship coefficient. Finally, we ran principal component analysis using TeraPCA^23^ to remove individuals who did not overlap with European samples in the 1000 Genomes dataset (mean PC value of top 6 PCs ± 3 SD). We ran further quality controls on the genotype data to remove individuals with missingness <0.98, SNPs with missingness > 0.98, minor allele frequency < 0.01 and Hardy-Weinberg threshold of 1e-06, resulting in a dataset of 330841 participants used for the analyses.

### PRS Calculation

PRScs software was used for the calculation of PRS^24^. This method utilizes a Bayesian regression framework and places a continuous shrinkage prior on the variant effect sizes using GWAS summary data and an external linkage disequilibrium (LD) panel. We used the 1000 Genomes European LD reference with HapMap3 SNPs, to update effect sizes jointly for all the SNPs in LD. The output is a file with updated effect size estimates which we used to calculate PRS using the score function in Plink. The summary statistics file required for the estimation of risk scores was obtained from the latest GWAS meta-analysis of TS^11^ with 6,133 cases and 13,565 controls to compute a weighted, mean score for each individual separately. The GWAS for other disorders were obtained from the Psychiatric Genomics Consortium^25-27^ and the number of SNPs used for PRS calculation are mentioned in the Supplementary Table 1.

### Phenome Wide Association Study

PHESANT package was used to run the PheWAS^28^. This tool was designed specifically to study the associations in UKB data. It classifies each phenotype as one of four data types: continuous, binary, ordered categorical, or unordered categorical and then estimates the association between PRS scores and each phenotype using a regression model that fits the data type of the phenotype (linear, logistic-binary, ordered-logistic, or multinomial-logistic). We included 2242 phenotypes which were classified into 90 UKB categories and were manually grouped into five categories in our analysis (biochemical markers, cognition and mental health, disease diagnosis, health and medical history, socio-demographics). The phenotypes in the disease diagnosis category were obtained after mapping the 17,000 ICD-10 diagnosis codes of the UKB to 1430 phenotypic codes using the Natural Language Processing (NLP) method implemented in the R package Phewas^29^. The number of phenotypes in each category are shown in Table 1 and the sub-categories are shown in Figure 1 and Supplementary Table 2. Age, sex, and genotyping batch derived from the UKB data and top 10 PCs calculated using TeraPCA were used as covariates for the analysis. To account for multiple testing, we used a Bonferroni threshold of p<2.3×10^−5^ (0.05/2242) for the results of our main analyses. Given the possible correlation between various phenotypes, a Bonferroni threshold could be considered to be overly conservative, so we also used a false discovery rate of 0.05 adjusted threshold (p <0.00495) to identify additional phenotypes associated with TS PRS in a secondary analysis. For the sex specific PheWAS analysis, we split the data into two parts (179976 females and 153440 males) based on the sex data provided in UKB and ran the analyses independently for both datasets.

**Table 1:** Total number of phenotypes analyzed by category.

| Categories | Total number |
| --- | --- |
| Biochemical Measures | 167 |
| Cognition and Mental Health | 223 |
| Disease Diagnoses | 1430 |
| Health and Medical History | 270 |
| Sociodemographics | 152 |

**Figure 1:**
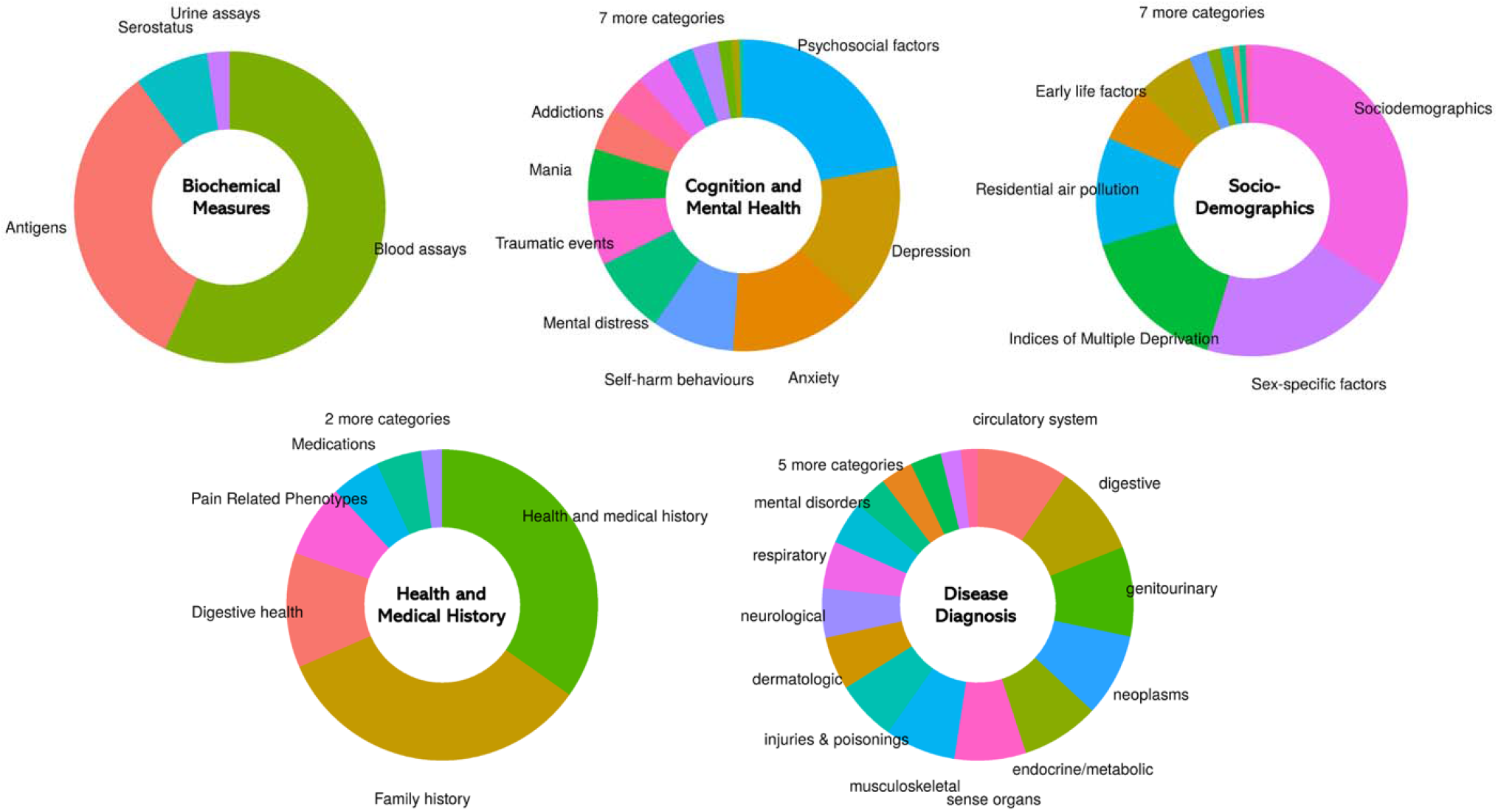
Overview of the phenotypic categories in UK Biobank with the size of pie chart sections indicating the number of included outcomes: biochemical measures (167), cognition and mental health (223), disease diagnosis (1430), health and medical history (270) and sociodemographics (152).

## Results

### Phenotypes associated with TS

The study included 330,841 unrelated individuals from the UKB dataset that were selected after filtering out individuals with non-European ancestry and other quality control measures. We tested the association of the scores with 2,242 phenotypes grouped into five major categories.

The overall results of our TS PRS-PheWAS are shown in Figure 2 and additional file 1. The analysis identified 57 outcomes significantly associated with TS PRS after Bonferroni correction for multiple testing (p<2.23×10^−5^) (Figure 3 and Supplementary Table 3a – 3e). The variable “Seen doctor for nerves, anxiety or depression” had the strongest association with TS PRS (beta: 0.031, p-value: 9.2×10^−17^). Twenty other cognition and mental health phenotypes were also observed to be significantly associated with TS PRS, with all of them having a positive association at the significant thresholds. This included associations for psychosocial factors like neuroticism score (beta: 0.026, p-value: 2.87×10^−15^), “worrier or anxious feelings” (beta: 0.026, p-value: 8.2×10^−13^), and the “tense/highly strung” trait (beta: 0.034, p-value:4.28×10^−13^). Various other behavioral traits were also found to be positively associated with TS PRS, including “increased mood swings” (beta: 0.019, p-value: 9.8×10^−8^), higher chance of “feeling miserable” (beta: 0.017, p-value: 9.32×10^−6^), and higher chance of “feeling unenthusiastic/disinterested for a whole week” (beta: 0.036, p-value: 3.05×10^−8^). We also observed that higher TS genetic risk was associated with “higher chances of being physically abused by family as child” (beta: 0.038, p-value:1.57×10^−6^).

**Figure 2:**
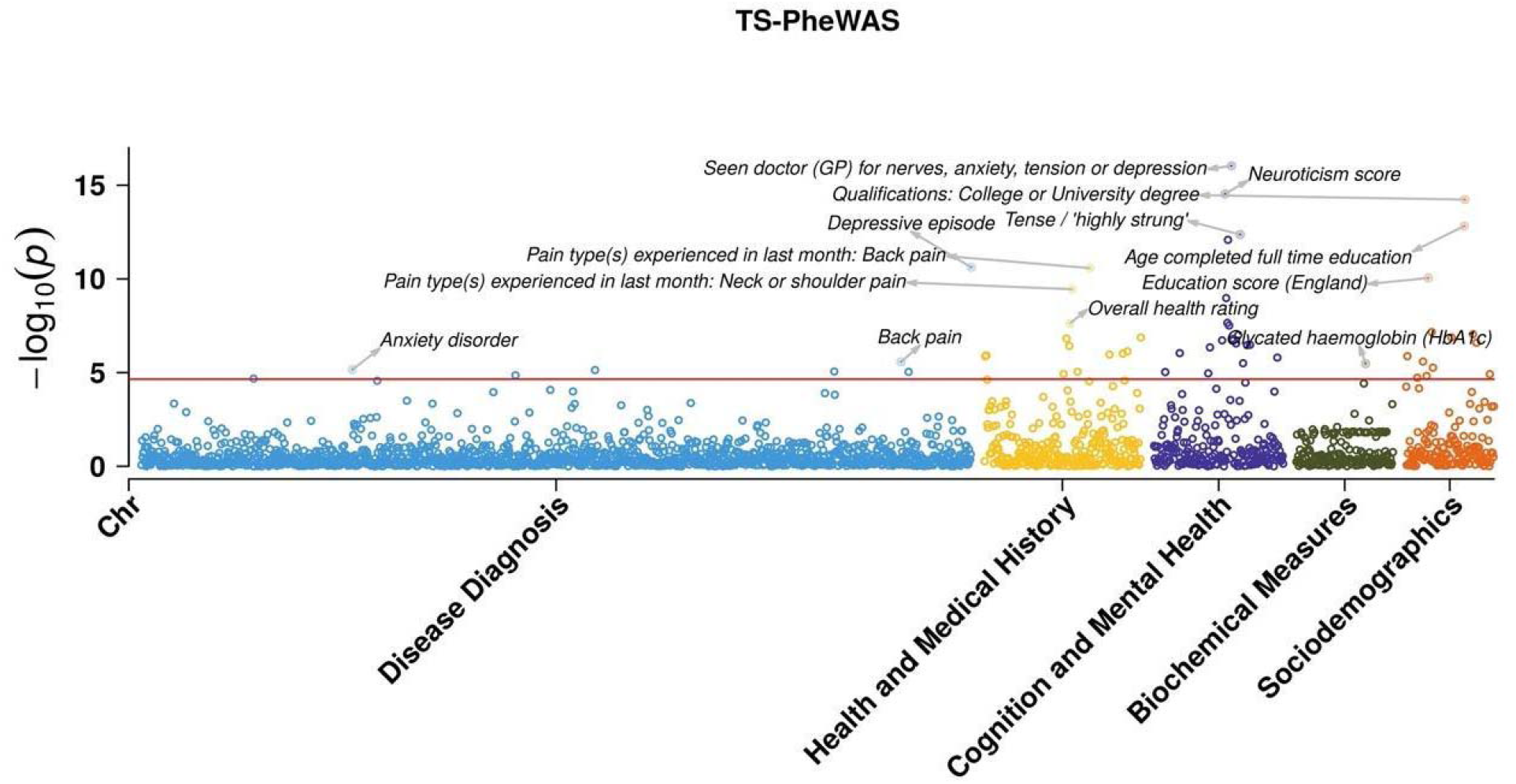
PheWAS Manhattan plot showing associations of phenotypes with TS PRS, grouped by categories. The horizontal line is marked at the Bonferroni threshold of significance for multiple testing (p<2.23×10^−5^). The top 3 significant associations in each category are highlighted and labeled.

**Figure 3:**
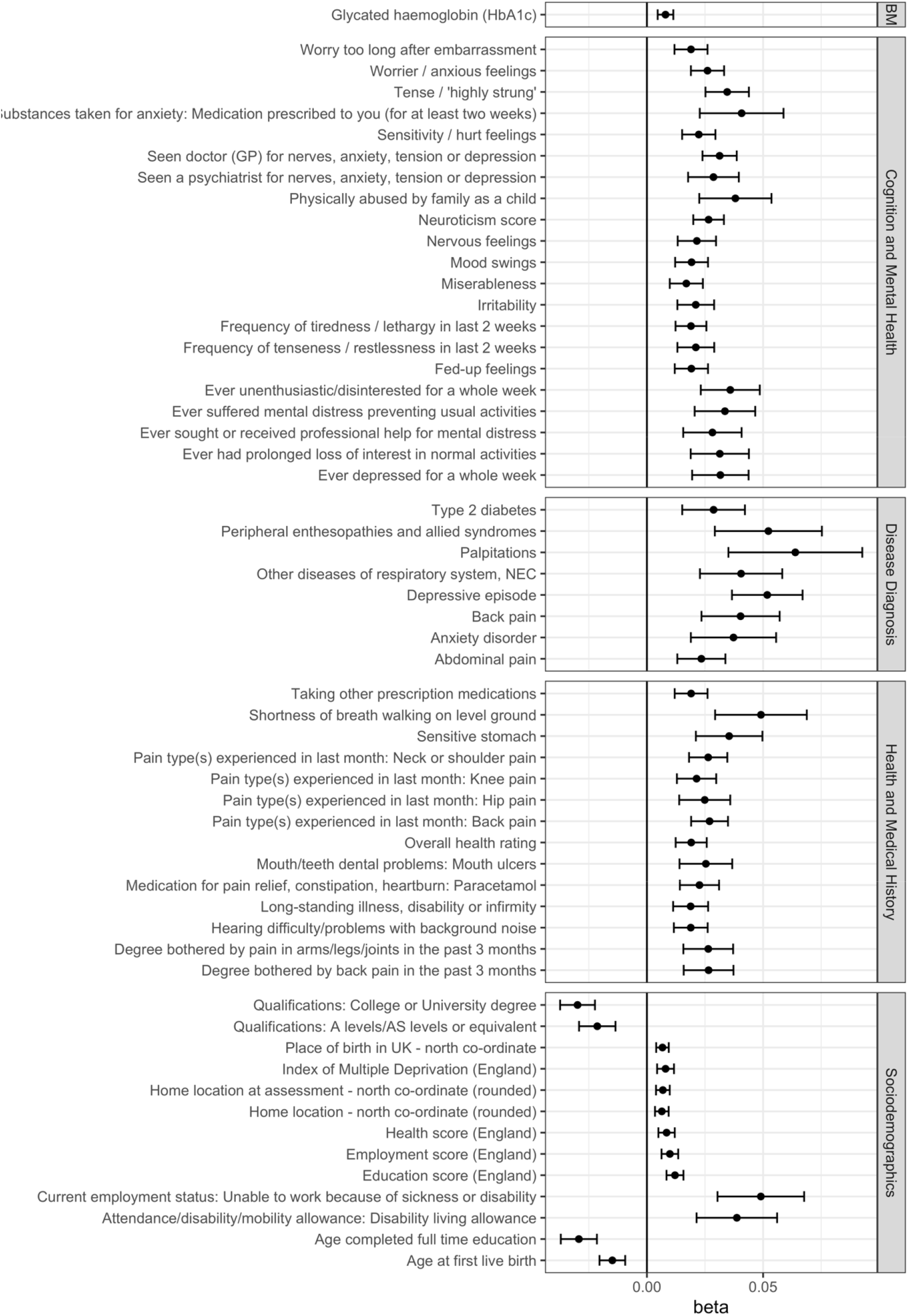
Forest plot showing phenotypes significantly associated with TS PRS, grouped by categories. BM: Biochemical Measures

Within the disease diagnosis category, we found eight outcomes significantly associated with TS risk scores with strong associations observed for psychiatric conditions including depressive episode (beta: 0.051, p-value: 2.38×10^−11^) and anxiety disorder (beta: 0.037, p-value: 6.9×10^−6^). We also observed non-mental health related diseases to be significantly associated with TS risk such as peripheral enthesopathies (beta: 0.052, p-value: 8.7×10^−6^), type 2 diabetes (beta: 0.028, p-value: 2.1×10^−5^), heart palpitations (beta: 0.063, p-value: 1.4×10^−5^), and diseases of the respiratory system (beta: 0.045, p-value: 1.2×10^−5^ – 2.5×10^−5^).

Fourteen health and medical history outcomes were observed to be significantly associated with TS risk, including several pain related phenotypes such as: back pain (beta: 0.027, p-value: 2.65×10^−11^), neck or shoulder pain (beta: 0.026, p-value: 3.54×10^−10^), and hip pain (beta: 0.025, p-value: 8.9×10^−6^). Other significant associations in this category included sensitive stomach (beta: 0.036, p-value: 1.3×10^−6^), shortness of breath walking on level ground (beta: 0.049, p-value:1.1×10^−6^), and mouth ulcers (beta: 0.025 p-value: 1.17×10^−5^). We also observed that individuals with a higher TS PRS had an increase in overall health rating (beta: 0.019, p-value: 2.35×10^−8^), indicating poorer overall self-reported health (as defined by UKB) correlated with a higher genetic risk of TS.

Among sociodemographic outcomes, 13 phenotypes were significantly associated with TS PRS, including parameters such as the index of multiple deprivation (beta: 0.008, p-value: 1.5×10^−5^), education score (beta: 0.012, p-value: 8.9×10^−11^), and employment score (beta: 0.01, p-value: 6.8×10^−8^). These results indicate that people with a higher TS PRS in the UKB data had a higher deprivation index and lower levels of education and employment. This was also highlighted by the negative association between TS PRS and the chance of having a college degree (beta: - 0.029, p-value: 5.8×10^−15^) or an A-level/AS-level qualification (beta: -0.021, p-value: 9×10^−8^). Finally, we also observed that people with a higher TS PRS had a lower age of first live birth (beta: -0.014, p-value: 1.3×10^−7^) and also completed their full-time education earlier (beta: -0.029, p-value: 1.5×10^−13^). Finally, in the biochemical measures category, glycated hemoglobin was found to have a positive association with TS PRS (beta: 0.007, p-value:3×10^−6^).

156 phenotypes were found to be significantly associated with TS PRS in our secondary analysis after FDR correction for multiple testing (Supplementary Figure 1). These included five biochemical measures, 41 cognition and mental health traits, 43 health and medical history outcomes, 25 socio-demographic measures, and 42 phenotypes from the disease diagnosis category. Some of the associations observed included complex medical conditions such as asthma, gastroesophageal reflux disease, irritable bowel syndrome, and hypercholesterolemia (beta: 0.02 - 0.035, p-value: 4.6×10^−3^-5.5×10^−4^). We also found fluid intelligence scores to be negatively associated with TS PRS (beta: -0.017, p-value: 3.7×10^−3^). Biochemical measures such as IGF1 and eosinophil count were also negatively associated with the risk of TS (beta: -0.005 to -0.008, p-value: 3.5×10^−3^ – 3.8×10^−5^).

### Comparison across neurodevelopmental disorders

We next proceeded to explore whether phenotypes associated with TS genetic risk were also associated with the genetic risk of other neurodevelopmental disorders that are known to be comorbid with TS, including ADHD, ASD, and OCD. The PRS scores of these disorders were weakly correlated with TS PRS (R2: 0.001 - 0.11). The PheWAS analysis was repeated for these disorders (additional file 2), and we compared the identified associations with TS following Bonferroni correction (Figure 3 and Table 2). For ADHD, we observed a total of 509 phenotypes significantly associated with ADHD PRS after correction for multiple testing, 53 of which were also found to be significantly associated with TS PRS. We observed that the phenotypic associations overlapping between ADHD and TS had stronger p-values and larger effect sizes in the same direction in relation to ADHD PRS compared to TS PRS. A total of 202 phenotypes were significantly associated with ASD PRS, and 34 of them were overlapping with TS phenotype associations. We observed that ASD PRS had a positive association with education-related phenotypes such as college qualifications (beta: 0.067, p-value:5.1×10^−69^) and A/AS level qualifications (beta: 0.051, p-value:3.6×10^−37^), which was contrary to what we found for TS PRS (which had a negative association). This suggests that people with higher genetic risk of ASD had an increased chance of completing college or school education. Among the 99 phenotypes significantly associated with the genetic risk of OCD, we observed that 26 overlapped with those associated with TS PRS. Similar to ASD, we found that education related phenotypes had a positive association with OCD PRS, again contrary to that seen with TS genetic risk. Additionally, this opposite direction of an association between OCD and TS was seen for all other phenotypes except for cognition and mental health traits. These included association with diseases such as Type 2 diabetes which had a positive association with TS and negative association with OCD risk, pain-related phenotypes, as well as indices of multiple deprivation such as the health score, education score, and employment scores. The associations between deprivation indices and OCD indicate that people with higher OCD PRS have lower deprivation scores in terms of health (beta: -0.012, p-value:2×10^−11^), education (beta: -0.018, p-value:1.7×10^−23^), and employment (beta: -0.013, p-value:3.7×10^−13^).

**Table 2:** Number of significant associations observed for the genetic risk of different neurodevelopmental disorders. The first column shows the different categories, and each column thereafter shows the number of significant associations observed for each of the disorders after multiple testing correction (p<2.23×10^−5^)

| Categories | Significant associations with TS PRS | Significant associations with ADHD PRS | Significant associations with ASD PRS | Significant associations with OCD PRS |
| --- | --- | --- | --- | --- |
| Biochemical Measures | 1 | 32 | 8 | 12 |
| Cognition and Mental Health | 21 | 105 | 71 | 13 |
| Disease Diagnosis | 8 | 196 | 1 | 2 |
| Health and Medical History | 14 | 125 | 26 | 4 |
| Sociodemographics | 13 | 83 | 34 | 15 |

Sixteen phenotypes were significantly associated with the genetic risk of all four disorders. Almost all of the cognition and mental health traits had similar directions of effect for the four disorders. Type 2 diabetes was significantly associated with all four disorders but was found to be positively associated with TS, ADHD, and ASD, but negatively associated with OCD. Overall, we observed a great degree of overlap between associations observed for TS and the different neurodevelopmental disorders.

### Sex-specific differences

To understand whether there were any differences in the associations between TS and the different phenotypes based on the sex of the individual, we subsequently performed sex specific PheWAS analysis (additional file 3). Figure 4 shows phenotypes significantly associated with TS PRS in males and females. Among females, we found 15 associations significantly associated with TS PRS, with the strongest association observed for college qualification (beta: -0.033, p-value: 1.7×10^−10^) (Supplementary Table 4). In males, we observed 18 associations significantly associated with TS genetic risk, with strong associations observed with mental health traits such as the “tense/highly strung” phenotype (beta: 0.047, p-value: 2.8×10^−10^) and neuroticism score (beta: 0.029, p-value: 5.3×10^−9^) (Supplementary Table 5). Although the direction of effect was similar, we observed a difference in the significance and strength of the association with various phenotypes. Overall, eight phenotypes were significantly associated with TS risk in both males and females, and these included socio-demographic traits and a few mental health outcomes. Notably, we observed that in the disease diagnosis category, only depression was significantly associated with TS risk in both sexes. In males, we observed significant associations with type 2 diabetes (beta: 0.038, p-value: 2×10^−5^) and palpitations (beta: 0.12, p-value: 1.4×10^−6^), both of which were not significantly associated in the PheWAS in females (p-value>0.05). Inversely, we also found that respiratory system diseases were significantly associated with TS genetic risk in females (beta: 0.061, p-value:2.5×10^−6^) but not in males.

**Figure 4:**
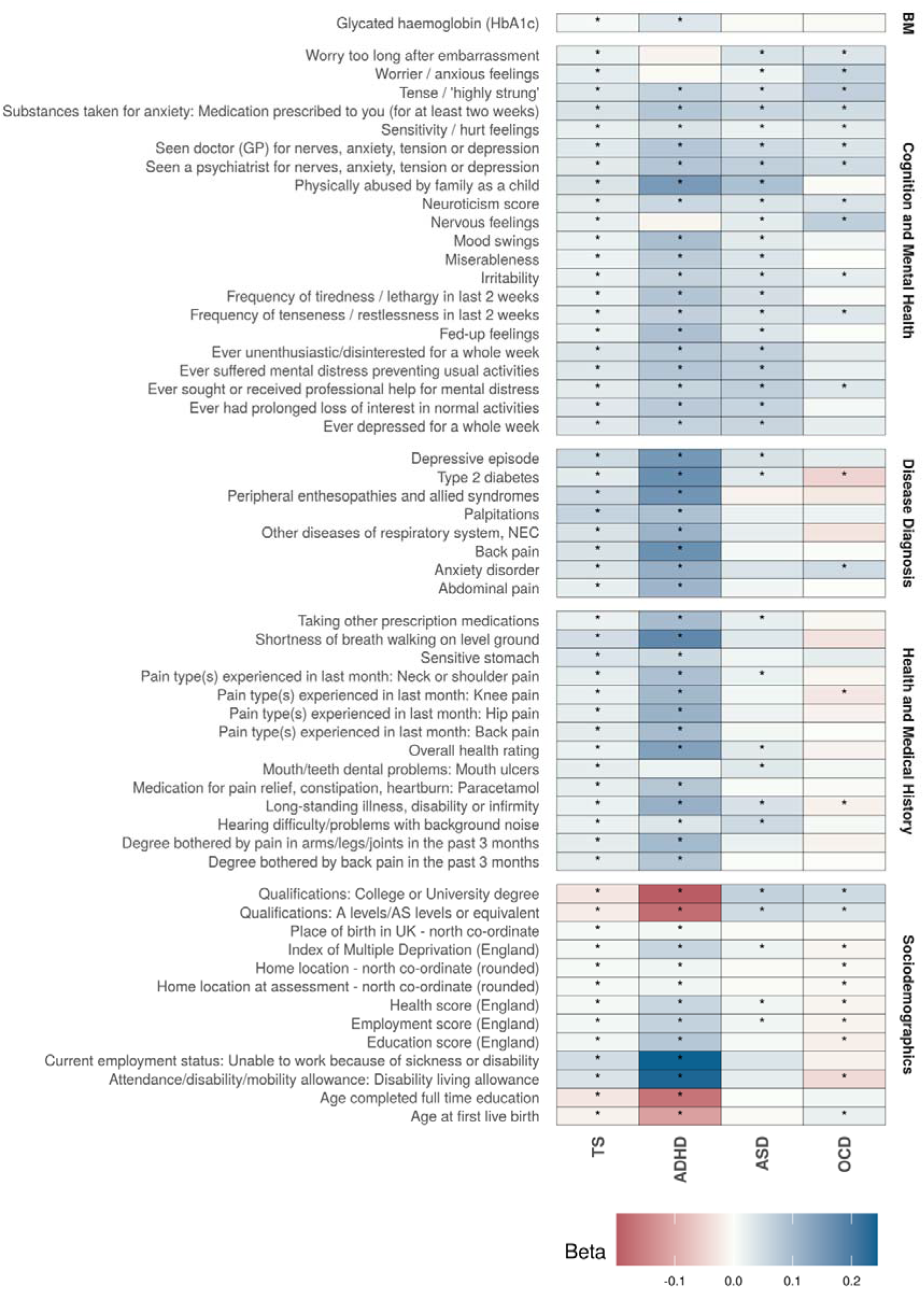
Cross–disorder comparison of significant associations with TS and at least one other disorder. (*) indicates significant association after multiple testing correction. BM: Biochemical Measures.

**Figure 5:**
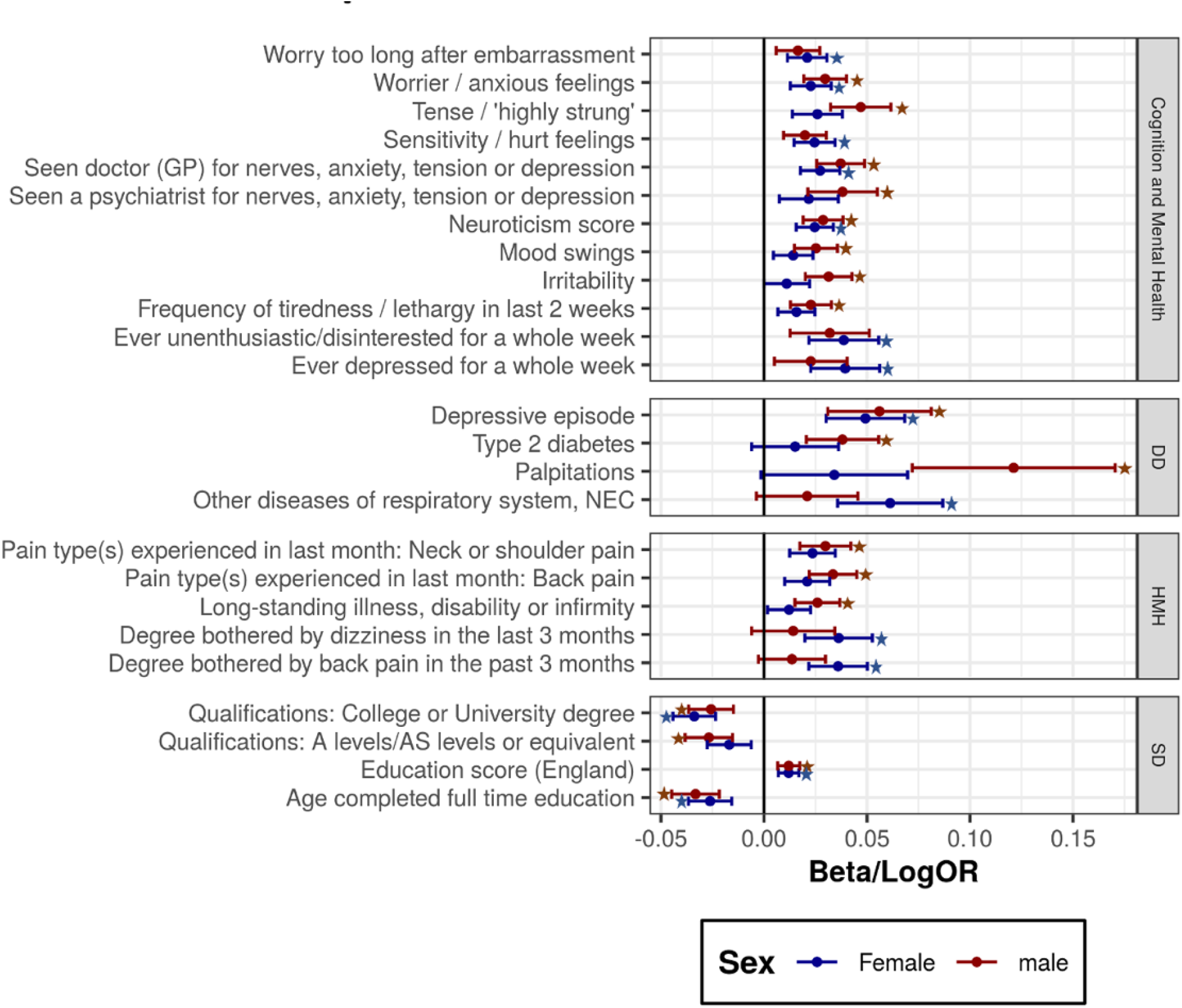
Phenotypes significantly associated with TS PRS in either males or females. (*) indicates significant after multiple testing correction. DD: Disease Diagnosis; HMH: Health and Medical History; SD: Socio-demographics.

## Discussion

We conducted a PRS-based PheWAS analysis to understand and identify associations between genetic liability for Tourette syndrome and 2242 phenotypes available in the UK Biobank dataset. Significant associations were observed with multiple general health, mental health, and sociodemographic traits. The strongest associations were found between TS PRS and mental health factors. Several associations with complex medical diseases were also observed, including associations with metabolic and respiratory disorders. Cross – disorder comparisons indicated that there was considerable overlap of phenotypes associated with the genetic risk of TS, ADHD, and ASD. Interestingly, OCD PRS had associations with a number of these same phenotypes albeit with an opposite direction of effect. Finally, sex specific PheWAS analysis highlighted difference in associations of complex disorders with TS PRS in males and females.

We were able to confirm previously observed associations of various phenotypes with TS, including neuroticism^30^, anxiety^31^, depression^32^, and lower educational attainment^33^. The association we observed between TS genetic risk and pain-related phenotypes was also previously observed in a prospective cohort study that investigated pain secondary to tics^34^. Additionally, epidemiological studies have also found a higher prevalence of type 2 diabetes and cardiovascular diseases among individuals with TS, in line with what we observed in our study, indicating that there might be some common biological mechanism between these disorders^12^. We found novel associations of TS genetic risk with various other mental health traits such as mood swings, irritability, nervous feelings, and loneliness that have shown to be associated with the genetic liability as well as clinical presentation of psychiatric disorders such as depression^35^. Interestingly we observed that higher genetic risk of TS was associated with lower maternal age of first live birth, which was also observed in epidemiological studies of ADHD^36^.

The results we obtained for the ADHD and ASD PheWAS were also consistent with previous phenome wide association studies for these disorders and we identified additional significant associations^18^. We observed that ASD and OCD had positive associations with school and college education, which has also been reported in genetic correlation analysis between these disorders and educational attainment^30^. This was also valid for the length of education, which had positive correlation with ASD and OCD, and negative correlation with TS and ADHD as seen both in the present and previous studies^30^. These results suggest that although these disorders are highly comorbid with TS and have strong genetic overlap, there are still considerable differences.

We also observed a difference in the association of outcomes with TS PRS based on the sex of the individuals. TS is more common in males than females and there are also differences in the associated phenotypes observed in each group^37-39^. We found differences primarily in associations with disorders such as type 2 diabetes and respiratory conditions, although no difference was observed in the overall PRS score distributions among the sexes. This suggests that there might be sex-chromosome differences or environmental factors influencing these dimorphisms^40^.

Although we identified strong associations with multiple traits using a PheWAS approach, there are certain limitations of this study. We used a large dataset of over 300,000 samples but there were only about 30 individuals with tics and stuttering and no individual with TS diagnosis (defined by ICD-10 diagnoses) even though the expected number would have been around 1800 (0.6%) based on the prevalence of the condition. Given the lack of TS cases, we could not calculate the exact predictive power of the TS PRS scores in the UKB dataset, which could have validated how well the PRS captured the differences between TS patients and controls.

Additionally, due to reduced overlap among the base and target datasets, the number of SNPs used for PRS calculation was lower for TS compared to the other neurodevelopmental disorders we studied here. This could possibly explain the low levels of correlation between TS PRS and the PRS of the other three disorders. We observe a higher number of associations for ADHD and ASD compared to TS and OCD, which could be attributed to the differences in the sample size of the discovery GWAS used for PRS calculation of these disorders^11,25-27^. Finally, our observed associations do not provide information about causality but should be followed up with additional study designs like mendelian randomization in appropriate datasets, or twin studies. We observed correlations with various social outcomes such as education and employment; but again, our results do not infer causality. Calculation of genetic risk estimates based on large-scale and more precise GWAS data are likely to be more predictive and may identify more outcomes associated with the disorder.

In conclusion, we showed that genetic liability of TS is associated with various phenotypic outcomes, several of which are also shared across other neurodevelopmental disorders like ADHD, ASD, and OCD. Phenotype associations with TS also appeared to differ based on sex. Our results suggest that it is important to consider a broad range of mental health, general health, and even sociodemographic outcomes associated with TS and other neurodevelopmental disorders to shed light in the complex etiology and related pathways underlying these conditions.

## Data Availability

All data produced in the present work are contained in the manuscript.

## Acknowledgments

The EMTICS collaborative group also includes Alan Apter, Juliane Ball, Benjamin Bodmer, Emese Bognar, Judith Buse, Marta Correa Vela, Carolin Fremer, Blanca Garcia-Delgar, Mariangela Gulisano, Annelieke Hagen, Julie Hagstrøm, Marcos Madruga-Garrido, Peter Nagy, Alessandra Pellico, Daphna Ruhrman, Jaana Schnell, Paola Rosaria Silvestri, Liselotte Skov, Tamar Steinberg, Friederike Tagwerker Gloor, Victoria L. Turner, Elif Weidinger

The TS-EUROTRAIN network also includes John Alexander, Tamas Aranyi, Wim R. Buisman, Jan K. Buitelaar, Nicole Driessen, Petros Drineas, Siyan Fan, Natalie J. Forde, Sarah Gerasch, Odile A. van den Heuvel, Cathrine Jespersgaard, Ahmad S. Kanaan, Harald E. Möller, Muhammad S. Nawaz, Ester Nespoli, Luca Pagliaroli, Geert Poelmans, Petra J. W. Pouwels, Francesca Rizzo, Dick J. Veltman, Ysbrand D. van der Werf, Joanna Widomska, Nuno R. Zilhäo

## Funding Statement

This work was funded by NIH grants R01NS105746, R01MH126213 and NSF grants 1715202, and 2006929. ZT was supported by Lundbeck Foundation (R100-A9332). YW was funded by the Deutsche Forschungsgemeinschaft. DB was supported by KNAW Academy Professor Award (PAH/6635). PJ and CZ were funded by the National Science Center, Poland: UMO-2016/23/B/NZ2/03030. BH is an employee of Boehringer Ingelheim Pharma. AS received support from the NIHR UCL/H Biomedical Research Centre. CM is supported by NIH grants (R01NS105746; R01NS102371)

## Conflict of Interests

The authors declare no conflict of interests.

## Additional File Legends

**Additional File 1**: Phenome Wide Associations of Phenotypes with TS PRS. Phenotypes arranged based on their main categories.

**Additional File 2**: Phenome Wide Associations of Phenotypes with PRS of other neurodevelopmental disorders. Each sheet represents results with each of the 3 disorders: ADHD, ASD, and OCD. Phenotypes arranged based on their main categories.

**Additional File 3**: Phenome Wide Associations of Phenotypes with TS PRS separated by sex. First sheet represents results of association with TS PRS in females and second for males. Phenotypes arranged based on their main categories.

## Notes

### Competing Interest Statement

The authors have declared no competing interest.

### Author Declarations

This research has been conducted using the UK Biobank Resource under the application number 61553

