## Supplementary for "Polygenic risk score-based phenome-wide association study identifies novel associations for Tourette syndrome"

Jain et al.

### List of tables


| | | |
|---|---|---|
| 1 | GWAS studies and Number of SNPs used for PRS calculation | 2 |
| 2 | Phenotype sub-categories | 3 |
| 3a | TS PheWAS significant associations – Biochemical Measures | 4 |
| 3b | TS PheWAS significant associations – Cognition and Mental Health | 4 |
| 3c | TS PheWAS significant associations – Disease Diagnosis | 5 |
| 3d | TS PheWAS significant associations – Health and Medical History | 5 |
| 3e | TS PheWAS significant associations – Sociodemographics | 6 |
| 4 | Significant associations – PheWAS female | 7 |
| 5 | Significant associations – PheWAS male | 8 |


### List of figures


| | | |
|---|---|---|
| 1 | Significant associations with TS PRS after FDR correction | 9 |

**References** 10

**Supplementary table 1**: GWAS studies used for PRS Calculations

| Disorder | Cases | Controls | N SNPs (PRS) | Reference |
|---|---|---|---|---|
| TS | 6,133 | 13,565 | 307,955 | (1) |
| ADHD | 20,183 | 35,191 | 924,303 | (2) |
| ASD | 18,381 | 27,969 | 958,504 | (3) |
| OCD | 2,688 | 7,037 | 964,437 | (4) |

**Supplementary table 2**: List of Sub-categories of phenotypes

| Category | Sub-Category | Number of Phenotypes |
|---:|---:|---:|
| Biochemical Measures | Antigens | 56 |
| Biochemical Measures | Blood assays | 96 |
| Biochemical Measures | Serostatus | 13 |
| Biochemical Measures | Urine assays | 4 |
| Cognition and Mental Health | Addictions | 10 |
| Cognition and Mental Health | Anxiety | 31 |
| Cognition and Mental Health | Depression | 34 |
| Cognition and Mental Health | Fluid intelligence and trail making | 10 |
| Cognition and Mental Health | Happiness and subjective well-being | 3 |
| Cognition and Mental Health | Mania | 12 |
| Cognition and Mental Health | Mental distress | 18 |
| Cognition and Mental Health | Pairs matching | 6 |
| Cognition and Mental Health | Psychosocial factors | 49 |
| Cognition and Mental Health | Self-harm behaviours | 19 |
| Cognition and Mental Health | Symbol digit substitution | 6 |
| Cognition and Mental Health | Traumatic events | 15 |
| Cognition and Mental Health | Unusual and psychotic experiences | 9 |
| Disease Diagnosis | circulatory system | 136 |
| Disease Diagnosis | congenital anomalies | 48 |
| Disease Diagnosis | dermatologic | 79 |
| Disease Diagnosis | digestive | 135 |
| Disease Diagnosis | endocrine/metabolic | 116 |
| Disease Diagnosis | genitourinary | 133 |
| Disease Diagnosis | hematopoietic | 46 |
| Disease Diagnosis | infectious diseases | 50 |
| Disease Diagnosis | injuries & poisonings | 90 |
| Disease Diagnosis | mental disorders | 64 |
| Disease Diagnosis | musculoskeletal | 105 |
| Disease Diagnosis | neoplasms | 123 |
| Disease Diagnosis | neurological | 73 |
| Disease Diagnosis | pregnancy complications | 30 |
| Disease Diagnosis | respiratory | 70 |
| Disease Diagnosis | sense organs | 106 |
| Disease Diagnosis | symptoms | 25 |
| Health and Medical History | Digestive health | 33 |
| Health and Medical History | Family history | 93 |
| Health and Medical History | Health and medical history | 102 |
| Health and Medical History | supplements and medication | 27 |
| Health and Medical History | Pain Related Phenotypes | 21 |
| Sociodemographics | Early life factors | 9 |
| Sociodemographics | Greenspace and coastal proximity | 9 |
| Sociodemographics | Home and work locations | 3 |
| Sociodemographics | Indices of Multiple Deprivation | 24 |
| Sociodemographics | Characteristics and reception | 4 |
| Sociodemographics | Residential air and noise pollution | 20 |
| Sociodemographics | Sex-specific factors | 31 |
| Sociodemographics | Sociodemographics | 52 |

**Supplementary table 3a**: Phenotypes significantly associated with TS PRS – Biochemical Measures category

| **Phenotype** | **beta** | **se** | **pvalue** |
|---|---|---|---|
| Glycated hemoglobin (HbA1c) | 0.007990481 | 0.001717972 | 3.30E-06 |

**Supplementary table 3b**: Phenotypes significantly associated with TS PRS – Cognition and Mental Health category

| **Phenotype** | **beta** | **se** | **pvalue** |
|---|---|---|---|
| Seen doctor (GP) for nerves, anxiety, tension or depression | 0.031276575 | 0.003761667 | 9.19E-17 |
| Neuroticism score | 0.026540103 | 0.003360914 | 2.87E-15 |
| Tense / 'highly strung' | 0.03453692 | 0.004766234 | 4.28E-13 |
| Worrier / anxious feelings | 0.026067196 | 0.003641953 | 8.21E-13 |
| Sensitivity / hurt feelings | 0.02231224 | 0.003657122 | 1.05E-09 |
| Frequency of tiredness / lethargy in last 2 weeks | 0.018950542 | 0.003388182 | 2.23E-08 |
| Ever unenthusiastic/disinterested for a whole week | 0.035871683 | 0.006477429 | 3.05E-08 |
| Mood swings | 0.019211779 | 0.003604395 | 9.81E-08 |
| Fed-up feelings | 0.019126261 | 0.003648398 | 1.58E-07 |
| Worry too long after embarrassment | 0.018965852 | 0.003628507 | 1.72E-07 |
| Irritability | 0.021010278 | 0.004033484 | 1.90E-07 |
| Frequency of tenseness / restlessness in last 2 weeks | 0.021037343 | 0.004042269 | 1.95E-07 |
| Seen a psychiatrist for nerves, anxiety, tension, or depression | 0.028632456 | 0.005571249 | 2.75E-07 |
| Nervous feelings | 0.021483792 | 0.004207695 | 3.29E-07 |
| Ever depressed for a whole week | 0.031597142 | 0.006198293 | 3.43E-07 |
| Ever suffered mental distress preventing usual activities | 0.0335722 | 0.006653632 | 4.51E-07 |
| Ever had prolonged loss of interest in normal activities | 0.031359828 | 0.006387983 | 9.13E-07 |
| Physically abused by family as a child | 0.038067792 | 0.007926903 | 1.57E-06 |
| Miserableness | 0.016949836 | 0.003637477 | 3.16E-06 |
| Substances taken for anxiety: Medication prescribed to you | 0.04075991 | 0.009197465 | 9.33E-06 |
| Ever sought or received professional help for mental distress | 0.028184137 | 0.00640976 | 1.10E-05 |

**Supplementary table 3c**: Phenotypes significantly associated with TS PRS – Disease Diagnosis category

| Phenotype | beta | se | pvalue |
|---|---:|---:|---:|
| Depressive episode | 0.051782103 | 0.007752179 | 2.38E-11 |
| Back pain | 0.04029887 | 0.00858208 | 2.65E-06 |
| Anxiety disorder | 0.037252578 | 0.0093597 | 6.89E-06 |
| Other diseases of respiratory system, NEC | 0.040522754 | 0.009037249 | 7.33E-06 |
| Peripheral enthesopathies and allied syndromes | 0.052264889 | 0.011754039 | 8.72E-06 |
| Abdominal pain | 0.023425516 | 0.005279084 | 9.10E-06 |
| Palpitations | 0.063867017 | 0.014697361 | 1.39E-05 |
| Type 2 diabetes | 0.028680425 | 0.006884423 | 2.10E-05 |

**Supplementary table 3d**: Phenotypes significantly associated with TS PRS – Health and Medical History category

| Phenotype | beta | se | pvalue |
|---|---:|---:|---:|
| Pain type(s) experienced in last month: Back pain | 0.026955263 | 0.004044663 | 2.65E-11 |
| Pain type(s) experienced in last month: Neck or shoulder pain | 0.026363456 | 0.00420298 | 3.55E-10 |
| Overall health rating | 0.019057118 | 0.003412573 | 2.35E-08 |
| Taking other prescription medications | 0.019017416 | 0.003605174 | 1.33E-07 |
| Medication for pain relief,heartburn: Paracetamol | 0.022608598 | 0.00430526 | 1.51E-07 |
| Hearing difficulty/problems with background noise | 0.018863865 | 0.003711667 | 3.73E-07 |
| Pain type(s) experienced in last month: Knee pain | 0.021360894 | 0.004312795 | 7.31E-07 |
| Long-standing illness, disability, or infirmity | 0.018801412 | 0.003838125 | 9.65E-07 |
| Shortness of breath walking on level ground | 0.049070123 | 0.010064553 | 1.08E-06 |
| Degree bothered by back pain in the past 3 months | 0.026532799 | 0.005464276 | 1.20E-06 |
| Sensitive stomach | 0.035381412 | 0.007307202 | 1.28E-06 |
| Degree bothered by pain in arms/leg in the past 3 months | 0.026394204 | 0.005462537 | 1.35E-06 |
| Pain type(s) experienced in last month: Hip pain | 0.024873744 | 0.005601627 | 8.98E-06 |
| Mouth/teeth dental problems: Mouth ulcers | 0.025362618 | 0.005787062 | 1.17E-05 |

**Supplementary table 3e**: Phenotypes significantly associated with TS PRS – Socio-demographics category

| **Phenotypes** | **beta** | **se** | **pvalue** |
|---|---:|---:|---:|
| Qualifications: College or University degree | -0.029863633 | 0.003824271 | 5.79E-15 |
| Age completed full time education | -0.029291899 | 0.003964888 | 1.50E-13 |
| Education score (England) | 0.012072247 | 0.001861634 | 8.91E-11 |
| Employment score (England) | 0.009867967 | 0.00182865 | 6.81E-08 |
| Qualifications: A levels/AS levels or equivalent | -0.021373731 | 0.003997793 | 8.99E-08 |
| Age at first live birth | -0.014886433 | 0.002822011 | 1.33E-07 |
| Unable to work because of sickness or disability | 0.049014332 | 0.009509013 | 2.54E-07 |
| Place of birth in UK - north co-ordinate | 0.006688758 | 0.001383233 | 1.33E-06 |
| Health score (England) | 0.00845566 | 0.00179795 | 2.57E-06 |
| Home location at assessment - north co-ordinate (rounded) | 0.006857597 | 0.001507999 | 5.43E-06 |
| Disability living allowance | 0.03869344 | 0.008843724 | 1.21E-05 |
| Index of Multiple Deprivation (England) | 0.008011607 | 0.00185223 | 0.000015236 |
| Home location - north co-ordinate (rounded) | 0.006414243 | 0.001498488 | 1.87E-05 |

**Supplementary table 4**: Phenotypes significantly associated with TS PRS in females

| **Phenotypes** | **beta** | **se** | **pvalue** |
|---|---|---|---|
| Sensitivity / hurt feelings | 0.024554381 | 0.005066822 | 1.26E-06 |
| Ever unenthusiastic/disinterested for a whole week | 0.038756164 | 0.008631537 | 7.10E-06 |
| Worry too long after embarrassment | 0.020998993 | 0.004907105 | 1.87E-05 |
| Seen doctor (GP) for nerves, anxiety, tension or depression | 0.027269906 | 0.004873705 | 2.20E-08 |
| Ever depressed for a whole week | 0.039459788 | 0.008545798 | 3.87E-06 |
| Worrier / anxious feelings | 0.022729625 | 0.005048433 | 6.72E-06 |
| Neuroticism score | 0.024657801 | 0.004602478 | 8.45E-08 |
| Other diseases of respiratory system, NEC | 0.061261966 | 0.013013583 | 2.50E-06 |
| Depressive episode | 0.049273606 | 0.009746161 | 4.28E-07 |
| Degree bothered by back pain in the past 3 months | 0.035989928 | 0.007246929 | 6.84E-07 |
| Degree bothered by dizziness in the last 3 months | 0.036256666 | 0.008350041 | 1.41E-05 |
| Age at first live birth | -0.01491267 | 0.002826986 | 1.33E-07 |
| Age completed full time education | -0.02613301 | 0.005352668 | 1.05E-06 |
| Qualifications: College or University degree | -0.03380712 | 0.005291744 | 1.68E-10 |
| Education score (England) | 0.011996824 | 0.002537246 | 2.27E-06 |

**Supplementary table 5**: Phenotypes significantly associated with TS PRS in males

| **Phenotypes** | **beta** | **se** | **pvalue** |
|---|---|---|---|
| Seen doctor (GP) for nerves, anxiety, tension, or depression | 0.037244529 | 0.005921185 | 3.17E-10 |
| Worrier / anxious feelings | 0.029714563 | 0.005260192 | 1.61E-08 |
| Neuroticism score | 0.02871147 | 0.004918909 | 5.33E-09 |
| Tense / 'highly strung' | 0.046998513 | 0.007451148 | 2.83E-10 |
| Seen a psychiatrist for nerves, anxiety, tension, or depression | 0.038160614 | 0.008612896 | 9.37E-06 |
| Frequency of tiredness / lethargy in last 2 weeks | 0.022795959 | 0.005055506 | 6.51E-06 |
| Irritability | 0.031414525 | 0.005761787 | 4.97E-08 |
| Mood swings | 0.025237894 | 0.005333735 | 2.22E-06 |
| Type 2 diabetes | 0.038116973 | 0.008950332 | 2.05E-05 |
| Palpitations | 0.121221502 | 0.025119212 | 1.39E-06 |
| Depressive episode | 0.056085476 | 0.012804602 | 1.19E-05 |
| Long-standing illness, disability, or infirmity | 0.025966399 | 0.005536675 | 2.73E-06 |
| Pain type(s) experienced: Back pain | 0.033509639 | 0.005886159 | 1.25E-08 |
| Pain type(s) experienced: Neck or shoulder pain | 0.02983633 | 0.006318959 | 2.34E-06 |
| Education score (England) | 0.012046066 | 0.002739844 | 1.10E-05 |
| Qualifications: A levels/AS levels or equivalent | -0.02673037 | 0.005882126 | 5.52E-06 |
| Qualifications: College or University degree | -0.02566017 | 0.005540527 | 3.64E-06 |
| Age completed full time education | -0.03319891 | 0.005903219 | 1.87E-08 |

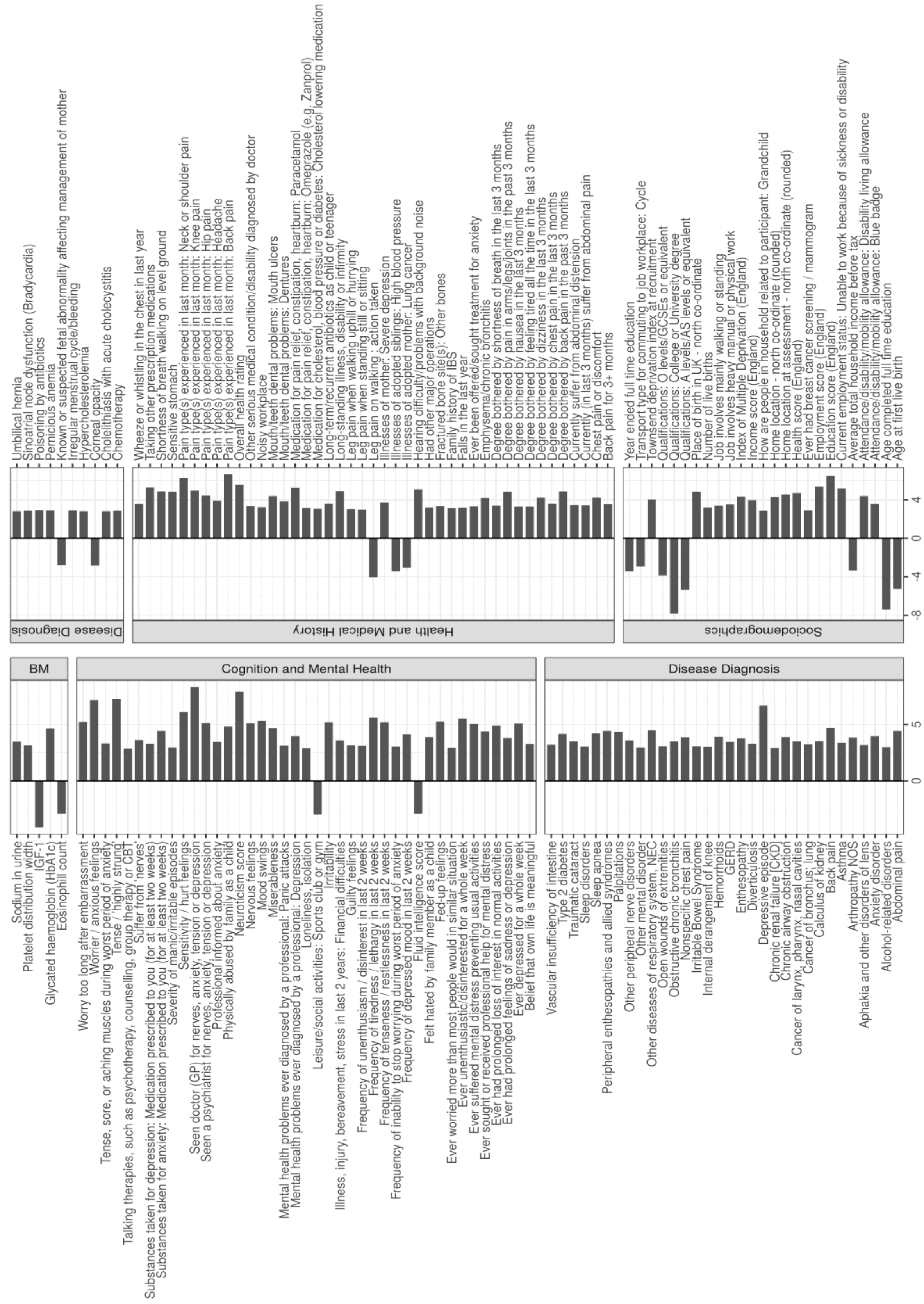


**Supplementary Figure 1:** Phenotypes Significantly associated with TS PRS after FDR correction arranged by categories. BM: Biochemical measures. Bars Indicate Z-score of association.
